# Localization-Aware Multiscale Deep Learning for Lumbar Foraminal Stenosis in Multi-Scanner Sagittal MRI: A Leakage-Controlled Evaluation

**DOI:** 10.64898/2026.09.02.26362033

**Authors:** Yousef Riyazifar

**Affiliations:** independent researcher in Karlsruhe, Germany

**Keywords:** Lumbar spine MRI, neural foraminal stenosis, deep learning, medical image segmentation, slice selection, object detection, ordinal classification, radiomics, uncertainty, explainable AI

## Abstract

Lumbar foraminal stenosis is a spatially localized and ordinal MRI interpretation problem: a useful computational system must first identify informative sagittal slices and foraminal regions before assigning severity. We present a retrospective, leakage-controlled multiscale deep-learning study using the public LSS MRI AISSLab cohort of 500 multi-scanner sagittal T2-weighted lumbar MRI examinations. A single frozen 70/15/15 patient partition was propagated across mid-sagittal anatomy segmentation, 2-D/2.5-D slice selection, anchor-free foraminal localization, four-grade region-of-interest (ROI) classification, radiomics, uncertainty analysis, and an exploratory whole-volume 3-D classifier. The selected U-Net achieved mean Dice 0.950 across five foreground anatomical labels (0.956 across all six released labels). The 2.5-D slice selector achieved held-out ROC AUC 0.926 (95% patient-clustered CI, 0.909–0.941). A threshold locked only on the tuning set yielded test sensitivity 0.876 (0.832– 0.919) and specificity 0.841 (0.813–0.869). Among 68 test patients with at least one annotated slice, an annotated slice appeared within the top three ranked slices in 68/68 patients (100%; exact 95% CI, 94.7–100%). The detector achieved localization AP50 0.530 but AP75 0.046, while 29.1% (26.0–32.0%) of annotation-negative test slices generated at least one prediction, identifying precise localization as the principal bottleneck. On expert-defined ROIs, a class-weighted scratch CNN achieved quadratic weighted kappa (QWK) 0.638 (0.550–0.706), with 92.7% (90.4–94.8%) of predictions within one grade. Moderate-or-worse and severe AUCs were 0.893 and 0.912, respectively. Compared with a 29- feature radiomics-SVM baseline, the CNN improved balanced accuracy by 0.205, macro-F1 by 0.173, and QWK by 0.360 using paired patient bootstrap. In a secondary uncertainty analysis, mean segmentation entropy strongly tracked mean surface error (Spearman *ρ* = 0.807, 95% CI 0.689–0.881). In contrast, the whole-volume 3-D CNN achieved AUC 0.639 (0.496–0.765) with poor calibration. The findings support an anatomically constrained, localization-aware strategy and demonstrate why raw accuracy or whole-volume classification alone can be misleading in highly imbalanced foraminal stenosis assessment.

## I. Introduction

**L** UMBAR neural foraminal stenosis is a common manifestation of degenerative lumbar disease and may contribute to radicular pain through loss of perineural space and nerve-root compromise. MRI is central to evaluation because it depicts intervertebral discs, neural structures, fat planes, posterior elements, and associated degenerative changes without ionizing radiation. Nevertheless, foraminal interpretation remains difficult because the clinically relevant anatomy occupies a small lateral region, appearance varies by level and slice position, and imaging severity is not perfectly synonymous with symptoms [1]–[3].

The four-grade Lee MRI system provides a reproducible framework for sagittal assessment. Grade 0 indicates no foraminal stenosis; grade 1 reflects partial perineural fat obliteration in two opposing directions; grade 2 reflects more extensive fat obliteration without nerve-root morphological change; and grade 3 reflects nerve-root collapse or morphological deformation [4]. The scale is therefore intrinsically *ordinal*. Errors between adjacent grades are not clinically equivalent to errors spanning the full severity range, which motivates weighted agreement, ordinal error, and clinically meaningful binary boundaries in addition to nominal accuracy.

Automated lumbar MRI interpretation has progressed from vertebral and disc localization to multi-task stenosis grading and evidence visualization [5]–[7] Hallinan *et al*., for example, demonstrated that a two-stage detection/classification approach can approach subspecialist agreement for dichotomized lumbar stenosis when trained on expert-labeled institutional MRI and externally tested [7]. Systematic reviews, however, continue to identify limited external testing, dataset heterogeneity, class imbalance, and insufficient reporting of clinically meaningful operating points as barriers to translation [8].

A particularly important methodological distinction is often obscured: *localized ROI grading is not the same task as finding the relevant anatomy in a complete MRI examination*. A network given an expert crop starts with most of the difficult spatial problem already solved. Conversely, a whole-volume classifier must discover small laterally positioned foramina, determine informative slices and levels, and then aggregate severity evidence. These tasks should not be treated as interchangeable measurements of “the same” model performance.

The LSS MRI AISSLab dataset now enables these questions to be studied on a public benchmark. The release contains 500 sagittal T2-weighted lumbar MRI examinations acquired across GE and Philips systems at 1.5 and 3 T, expert-verified foraminal bounding boxes with Lee-style severity grades, and expert-refined mid-sagittal segmentation masks [9], [10]. The associated data descriptor reported a complete CAD pipeline, while a subsequent 2026 study investigated ordinal grading using expert-localized ROIs and morphometric fusion; that study explicitly did not evaluate automated localization within the complete sagittal examination [11]. These works establish a strong benchmark but leave room for a controlled analysis of *where* information is gained or lost across spatial scales.

This study therefore asks a focused question: *how does performance change when lumbar MRI analysis progresses from anatomy-constrained representations to increasingly unconstrained spatial representations under one frozen patient split?* Rather than optimizing a single headline classifier, we evaluate the complete chain of anatomical segmentation, slice ranking, object localization, localized severity grading, radiomics, uncertainty, and whole-volume classification. The design emphasizes leakage control, validation-locked thresholds, patient-clustered confidence intervals, explicit failure analysis, and reproducibility in accordance with CLAIM 2024 reporting principles [12].

The primary contributions are:

1. a single patient-level partition reused across multiple MRI tasks, preventing cross-task slice or ROI leakage;
2. a validation-locked 2.5-D slice-selection operating point and patient-level top-*k* retrieval analysis;
3. an explicit decomposition of localization quality into AP50/AP75, class-aware detection, and prediction burden on annotation-negative slices;
4. ordinal and class-balanced evaluation of expert-localized severity classification, including paired patient-bootstrap comparison against radiomics;
5. an exploratory whole-volume 3-D comparator that is retained despite weak performance, avoiding selective reporting; and
6. a quantitative link between segmentation predictive entropy and patient-level surface error, complementing qualitative medical-image examples.

## II. Clinical and Technical Context

### A. Clinical Interpretation of Foraminal Stenosis

The neural foramen is bounded by pedicles, vertebral bodies, discs, and facet-related posterior structures. Loss of perineural intraforaminal fat and nerve-root deformation are among the most accepted MRI indicators of foraminal compromise [1]. The Lee system has shown high reproducibility in its original report [4], and later work found substantial correlation with clinical manifestations while also highlighting imperfect concordance between imaging grade and symptoms [2]. More recent high-resolution MRI work has proposed finer six-point systems, illustrating that clinically relevant foraminal morphology can be more nuanced than a simple four-class label [3]. Consequently, the current study treats the released four-grade reference as an imaging reference standard rather than as a direct surrogate for symptoms, disability, or surgical indication.

### B. Why Spatial Granularity Matters for AI

Automated stenosis systems may operate at several scales: full examination, selected slice, anatomically localized level, or expert-defined ROI. DeepSPINE and SpineNet demonstrated the value of explicit localization and multi-task modeling in lumbar MRI [5], [6]. The AISSLab data descriptor similarly decomposed analysis into slice selection, ROI detection, and severity classification [9]. The present study extends that decomposition by quantifying performance and failure modes at each spatial scale under one patient split and by adding a deliberately unconstrained 3-D comparator.

## III. Materials and Methods

### A. Study Design and Reporting Framework

This was a retrospective secondary computational study of de-identified public MRI data. No prospective clinical decision was made, no patient was recruited by the author, and no study prediction influenced care. Reporting terminology follows CLAIM 2024: “reference standard” is used instead of “ground truth,” the internal development subset used for model optimization is called the tuning/validation set, and the held-out subset is called the internal test set [12].

The complete public cohort was used; no formal samplesize calculation was performed because the available cohort size and annotation count were fixed by the public release. Quantitative test endpoints were reported with patient-level or patient-clustered uncertainty wherever possible. Age and sex were not part of the frozen analysis manifest and were not used as model inputs; demographic subgroup analysis was therefore not performed.

### B. Dataset, Reference Standard, and Ethics

The LSS MRI AISSLab dataset contains 500 unique patients with routine sagittal T2-weighted lumbar MRI acquired at Fırat University Hospital on Philips Ingenia 1.5 T/3 T, GE Signa HDxt 1.5 T, and GE Signa Excite 1.5 T systems [9]. The source release contains full DICOM series, mid-sagittal segmentation images/masks, and sagittal PNG images with PASCAL-VOC-style foraminal XML annotations. The dataset article reports 8,500 sagittal MRI slices and 2,979 expert-verified bounding-box annotations. The released analysis PNG manifest used here contained 6,644 slices. One box failed geometric validity checks, leaving 2,978 valid ROI annotations.

The reference standard provides laterality, lumbar level L1–L2 through L5–S1, bounding-box coordinates, and severity grade. The dataset article describes annotation and validation by expert neurosurgeons and reports institutional ethics approval (Fırat University Non-Interventional Research Ethics Committee, 2023/12-20, 14 September 2023), deidentification, and public release for research [9]. This study performed secondary analysis of the public de-identified resource and introduced no new human-subject interaction. The 2,978 valid ROIs comprised 2,009 grade-0 (normal), 507 grade-1 (mild), 254 grade-2 (moderate), and 208 grade-3 (severe) annotations. The imbalance is clinically and statistically important: 67.5% of all annotated ROIs were grade 0.

### C. Frozen Patient-Level Partition and Leakage Control

A deterministic patient split was created once and reused across tasks: 350 patients (70%) for training, 75 (15%) for tuning/validation, and 75 (15%) for internal testing. All slices, masks, and ROIs inherited their patient’s split. Thus, no slice or ROI from a test patient could enter training or architecture selection.

**Fig. 1.**
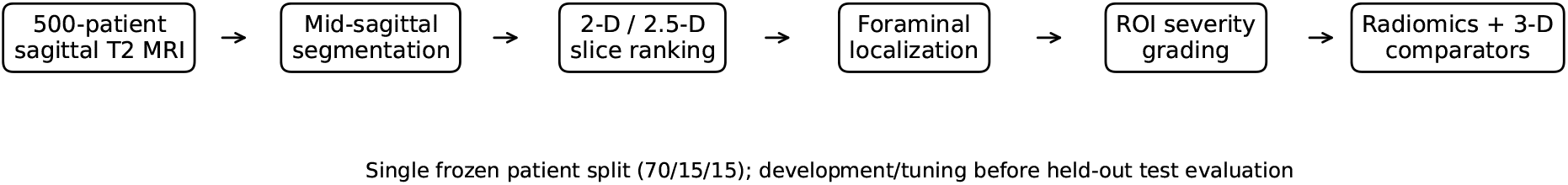
Study design. One frozen patient partition was propagated across anatomical segmentation, 2-D/2.5-D slice ranking, foraminal localization, expert-ROI severity grading, radiomics, uncertainty analysis, and an exploratory 3-D patient-level comparator. Model and threshold selection used only development/tuning data. Held-out test predictions were subsequently subjected to frozen postprocessing without retraining.

**Fig. 2.**
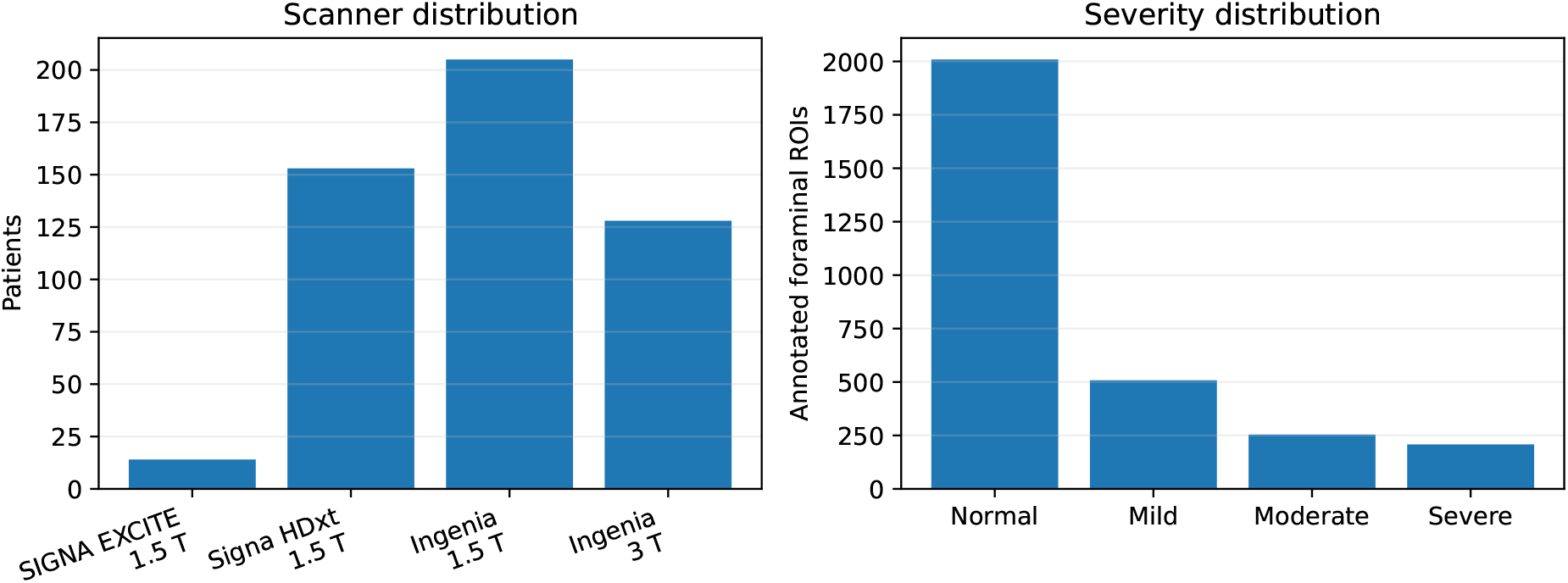
Cohort composition. Left: multi-scanner distribution across GE and Philips platforms at 1.5 and 3 T. Right: pronounced four-grade imbalance of valid foraminal annotations after geometric quality control.

Only 469 patients contained foraminal annotations, and one invalid ROI was excluded. The severity and 3-D experiments therefore contained 468 eligible patients: 329 training, 71 tuning, and 68 testing. ROI counts were 2,109/405/464. Slice selection used 4,640/1,006/998 slices across the same 350/75/75 patient partition. The held-out detector evaluation contained 225 annotation-bearing test slices from 68 patients and 773 annotation-negative slices from all 75 test patients.

Software-level stage and test-evaluation locks were used during the final run so completed test evaluation could not silently become part of subsequent model selection. Publication postprocessing operated only on frozen predictions, saved metrics, and frozen models; it did not retrain networks or optimize thresholds on test data.

### D. Image Normalization and General Preprocessing

Input images were converted to floating point; non-finite values were set to finite background values; and robust intensity normalization used percentile clipping followed by scaling to [0, 1]. Images were resized only after normalization to the task-specific input dimensions. Development-time augmentation, where applied, was restricted to the training partition. Tuning and test inference were deterministic.

### E. Mid-Sagittal Anatomical Segmentation

The public dataset provides one mid-sagittal MRI and a six-value expert-refined mask for each patient, representing the anterior/background region plus vertebral bodies, intervertebral discs, sacrum, Posterior A, and Posterior B [9]. The final result archive stores classes by grayscale mask code (0, 50, 100, 150, 200, 255), so quantitative class results are reported by released label index rather than imposing an unverified code-to-name mapping.

Standard U-Net [13], residual U-Net based on residual learning [14], and Attention U-Net [15] were trained on 320 *×* 320 images. Architecture selection used tuning-set mean Dice with HD95 as a complementary surface-quality criterion. The selected architecture was evaluated once on 75 internal-test patients. For class *c*, Dice was

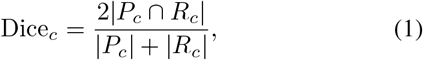

where *P*_*c*_ and *R*_*c*_ denote predicted and reference pixels. IoU, sensitivity, precision, HD95, and average symmetric surface distance (ASSD) were also calculated. Patient bootstrap confidence intervals used 2,000 resamples in the original run.

### F. 2-D and 2.5-D Slice Ranking

The slice-selection target was binary: whether a sagittal PNG had an associated XML foraminal annotation. This target denotes an *annotation-bearing/informative slice*; it is not a disease label because normal foramina can also be annotated. A 2-D CNN used one slice, whereas a 2.5-D CNN stacked the immediately preceding, current, and immediately following sagittal slices as three channels, with edge replication at volume boundaries. Inputs were resized to 224 *×* 224.

Model selection used tuning AUC. After the 2.5-D model was frozen, a clinically useful threshold was selected on the tuning set by requiring sensitivity ≥ 0.90 and maximizing specificity subject to that constraint. The resulting threshold (0.114394) was applied unchanged to the test set. Patient-cluster bootstrap confidence intervals used 5,000 resamples.

A second operational analysis ranked all test slices within each patient by frozen 2.5-D score. Among patients with at least one annotation-bearing slice, top-*k* hit rate was defined as the fraction for whom at least one reference-positive slice appeared among the *k* highest-scoring slices. Exact Clopper– Pearson 95% binomial intervals were used for top-*k* proportions because top-3 achieved 68/68 hits and a percentile bootstrap would otherwise be degenerate.

### G. Anchor-Free Foraminal Localization

A custom fully convolutional anchor-free grid detector operated on 320 *×* 320 sagittal images. Each grid location predicted objectness, four severity-class logits, and normalized bounding-box parameters. Training combined objectness, box-regression, and classification losses. Frozen inference used a score threshold of 0.25 and non-maximum suppression at IoU 0.4.

On annotation-bearing test slices, predictions were sorted by confidence and matched one-to-one with reference boxes. Localization AP was computed at IoU 0.50 and 0.75. Class-aware AP additionally required the predicted severity grade to match the reference grade. To characterize prediction burden outside the annotated-slice subset, the same frozen detector and thresholds were applied to all 773 test slices with no XML annotation. These are termed *annotation-negative*, not clinically normal, because absence of an XML file does not prove absence of subtle pathology.

### H. Expert-ROI Four-Grade Severity Classification

Each valid expert bounding box was enlarged by a 20% contextual margin, clipped to image bounds, normalized, and resized to 224 *×* 224. Two candidates were developed: a compact grayscale CNN trained from scratch and SqueezeNet transfer learning [16]. Inverse-frequency class weighting mitigated dominance of grade 0. Model selection prioritized macro-F1 and balanced accuracy rather than accuracy alone.

The frozen scratch CNN was tested on 464 ROIs from 68 patients. Metrics included accuracy, balanced accuracy, macro-F1, mean absolute ordinal error (MAE), quadratic weighted kappa (QWK), and within-one-grade agreement. Because the output probabilities retain ordinal information, three clinically interpretable binary scores were also formed: any stenosis (*P*_1_ + *P*_2_ + *P*_3_), moderate-or-worse (*P*_2_ + *P*_3_), and severe (*P*_3_). Patient-clustered ROC AUC intervals used 5,000 bootstrap resamples.

QWK was calculated with quadratic disagreement weights

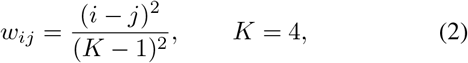

so errors spanning multiple grades receive greater penalty than adjacent-grade errors.

### I. Radiomics-SVM Comparator

To test whether handcrafted descriptors could explain the same ordinal signal, 29 features were extracted from exactly the same expert-defined ROIs. Feature families included first-order intensity statistics, gray-level co-occurrence texture, local binary pattern descriptors, histogram-of-oriented-gradient information, Laplacian-of-Gaussian responses, and gradient statistics. Standardization parameters were learned on development data only. PCA dimensionality (nominal grid values 16, 32, 64) and RBF-SVM box constraint (0.1, 1, 10) were optimized by grouped five-fold development cross-validation using macro-F1. Because only 29 source features existed, the final PCA transform correctly retained a maximum of 29 dimensions even though the best nominal grid label was 64. The frozen radiomics model was evaluated on the same 464 test ROIs, enabling patient-clustered paired comparison with the CNN.

### J. Whole-Volume 3-D Comparator

To test whether explicit localization could be omitted, sagittal image stacks from the 468 annotated patients were normalized and resampled to 128 *×* 128 *×* 16. A compact 3-D CNN predicted whether a patient had at least one moderate or severe (≥ 2) annotation. The internal test set contained 68 patients: 35 positive and 33 negative. ROC AUC was the principal discriminator; fixed-threshold sensitivity/specificity, Brier score, expected calibration error (ECE), and scanner-group summaries were treated as descriptive exploratory end-points.

### K. Explainability and Segmentation Uncertainty

Occlusion sensitivity was generated for selected severity-classification ROIs by measuring the change in class score when local image regions were obscured. These visualizations were used qualitatively only and were not treated as proof of causal model reasoning.

For segmentation, saved softmax maps enabled per-pixel Shannon entropy

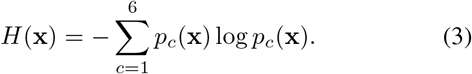

Mean entropy and the 95th percentile of entropy were summarized per test patient. To test whether this uncertainty measure was informative rather than decorative, patient mean entropy was correlated post hoc with patient-mean Dice, HD95, and ASSD using Spearman rank correlation. Confidence intervals used 5,000 patient resamples. This is a secondary analysis and is interpreted as an association with segmentation error, not as calibrated epistemic uncertainty.

### L. Statistical Analysis

All inferential resampling respected patient clustering. For slice-level and ROI-level metrics, a sampled patient contributed all of that patient’s slices or ROIs. For paired CNN-versus-radiomics comparisons, the same sampled patients were used for both methods. Confidence intervals were percentile 95% intervals unless otherwise stated. We emphasize effect sizes and intervals rather than relying on isolated *p* values. Secondary subgroup results by level, side, or scanner were descriptive because subgroup sizes were not powered a priori.

## IV. Results

### A. Cohort Audit, Scanner Heterogeneity, and Label Imbalance

All 500 public patients had readable DICOM metadata in the final audit. Scanner groups were GE SIGNA EXCITE 1.5 T (*n* = 14), GE Signa HDxt 1.5 T (*n* = 153), Philips Ingenia 1.5 T (*n* = 205), and Philips Ingenia 3 T (*n* = 128). Thus, 372/500 examinations were acquired at 1.5 T and 128/500 at 3 T. Foraminal annotations were available for 469 patients; 31 patients had no released foraminal annotations because the source study considered the foramina inadequately visualized [9].

The valid 2,978-ROI distribution was strongly imbalanced: 67.5% normal, 17.0% mild, 8.5% moderate, and 7.0% severe. L3–L4 contributed the greatest number of annotated ROIs (795), whereas L5–S1 contributed the fewest (278). The imbalance motivated class-weighted learning and class-balanced/ordinal evaluation.

### B. Anatomical Segmentation Was Highly Accurate but Not Uniformly Perfect

On tuning data, standard U-Net achieved mean Dice 0.958 and mean HD95 14.8 pixels; Attention U-Net achieved Dice 0.957 but HD95 39.4 pixels; residual U-Net achieved Dice 0.944 and HD95 27.5 pixels. Standard U-Net was therefore selected.

Held-out Dice values across released mask labels 1–6 were 0.985, 0.981, 0.974, 0.954, 0.932, and 0.911, respectively (Table III). The six-label macro-average was 0.956; excluding the anterior/background label yielded foreground macro Dice 0.950. Mean foreground overlap was therefore high, but surface distances exposed localized outliers: the class-wise macro mean HD95 across all labels was 19.0 pixels, driven largely by labels 1 and 3. This is why segmentation is described as strong in bulk overlap rather than “perfect.”

**Table I.** Context Relative to Representative Lumbar MRI AI Studies.

| Study | Cohort | MRI input | Principal task | Relevance to present study |
| --- | --- | --- | --- | --- |
| SpineNet [5] | Institutional | Lumbar MRI | Automated classification and evidence visualization | Established deep-learning analysis of degenerative spinal MRI. |
| DeepSPINE [6] | 4,075 patients | Multi-series lumbar MRI | Vertebral segmentation, level designation, stenosis grading | Demonstrated value of localization and multi-input grading. |
| Hallinan *et al.* [7] | 446 internal + 100 external | Sagittal T1 + axial T2 | Two-stage detection/classification of central, lateral recess, and foraminal stenosis | Strong clinical benchmark with external testing and expert reader comparison. |
| AISSLab data descriptor [9] | 500 public patients | Sagittal T2 | Dataset + CAD technical validation | Source dataset; establishes expert annotations and public benchmark. |
| Phadke *et al.* [11] | 469 public patients | Expert-cropped sagittal T2 ROIs | Ordinal ROI grading + morphometry | Same public resource; automated full-exam localization not evaluated. |
| Present study | 500 public patients | Sagittal T2 | Multiscale segmentation, ranking, localization, ROI grading, radiomics, uncertainty, 3-D comparator | Focuses on leakage-controlled spatial granularity and failure analysis rather than a single classifier. |

**TABLE II.** Frozen Study Partition and Test Units.

| Task | Train patients | Tuning patients | Test patients | Test units | Test target |
| --- | --- | --- | --- | --- | --- |
| Mid-sagittal segmentation | 350 | 75 | 75 | 75 images × 6 labels | Anatomical mask overlap |
| 2-D/2.5-D slice selection | 350 | 75 | 75 | 998 slices | XML annotation presence |
| Foraminal detector | – | – | 68 | 225 annotated slices | Bounding-box localization |
| Detector negative-slice audit | – | – | 75 | 773 slices | Prediction burden without XML |
| Four-grade ROI classifier | 329 | 71 | 68 | 464 ROIs | Grade 0/1/2/3 |
| Radiomics comparator | 329 | 71 | 68 | Same 464 ROIs | Grade 0/1/2/3 |
| 3-D patient classifier | 329 | 71 | 68 | 68 volumes | Any grade $\geq 2$ annotation |

**TABLE III.** Held-Out Segmentation Performance by Released Mask Label.

| Label | Dice | 95% CI | IoU | Sens. | Prec. |
| --- | --- | --- | --- | --- | --- |
| 1 | 0.985 | 0.982–0.987 | 0.970 | 0.992 | 0.978 |
| 2 | 0.981 | 0.978–0.983 | 0.962 | 0.977 | 0.985 |
| 3 | 0.974 | 0.967–0.980 | 0.951 | 0.963 | 0.988 |
| 4 | 0.954 | 0.948–0.957 | 0.912 | 0.966 | 0.942 |
| 5 | 0.932 | 0.928–0.937 | 0.874 | 0.925 | 0.941 |
| 6 | 0.911 | 0.894–0.926 | 0.843 | 0.910 | 0.919 |

Crucially for an imaging manuscript, the quantitative values were reflected in the images themselves. Figure 4 shows a high-overlap case and the lowest patient-mean-Dice test case, selected objectively from the frozen metric table rather than by visual preference. Even in the challenging case, the main vertebral/disc contour is retained, while local segmentation defects explain the lower surface-based performance.

**Fig. 3.**
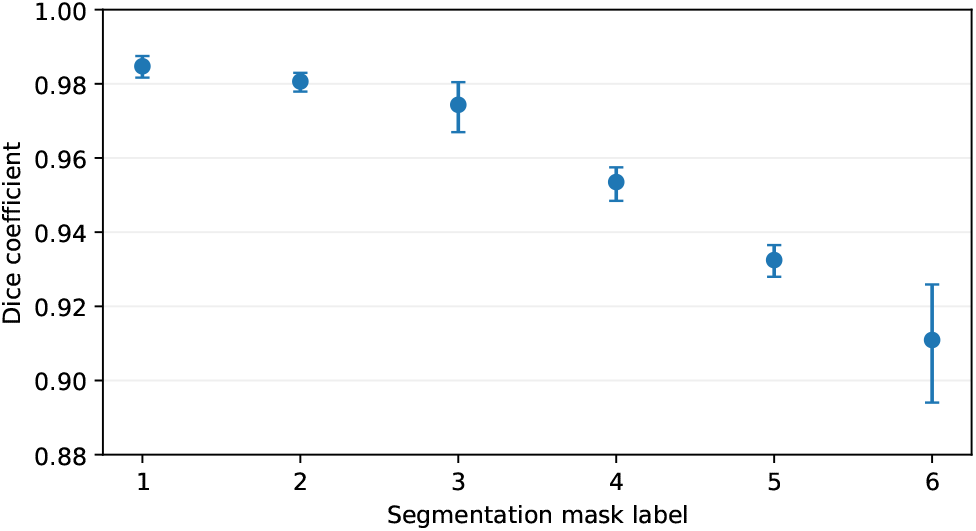
Held-out segmentation Dice and 95% patient-bootstrap confidence intervals for six released mask labels.

**Fig. 4.**
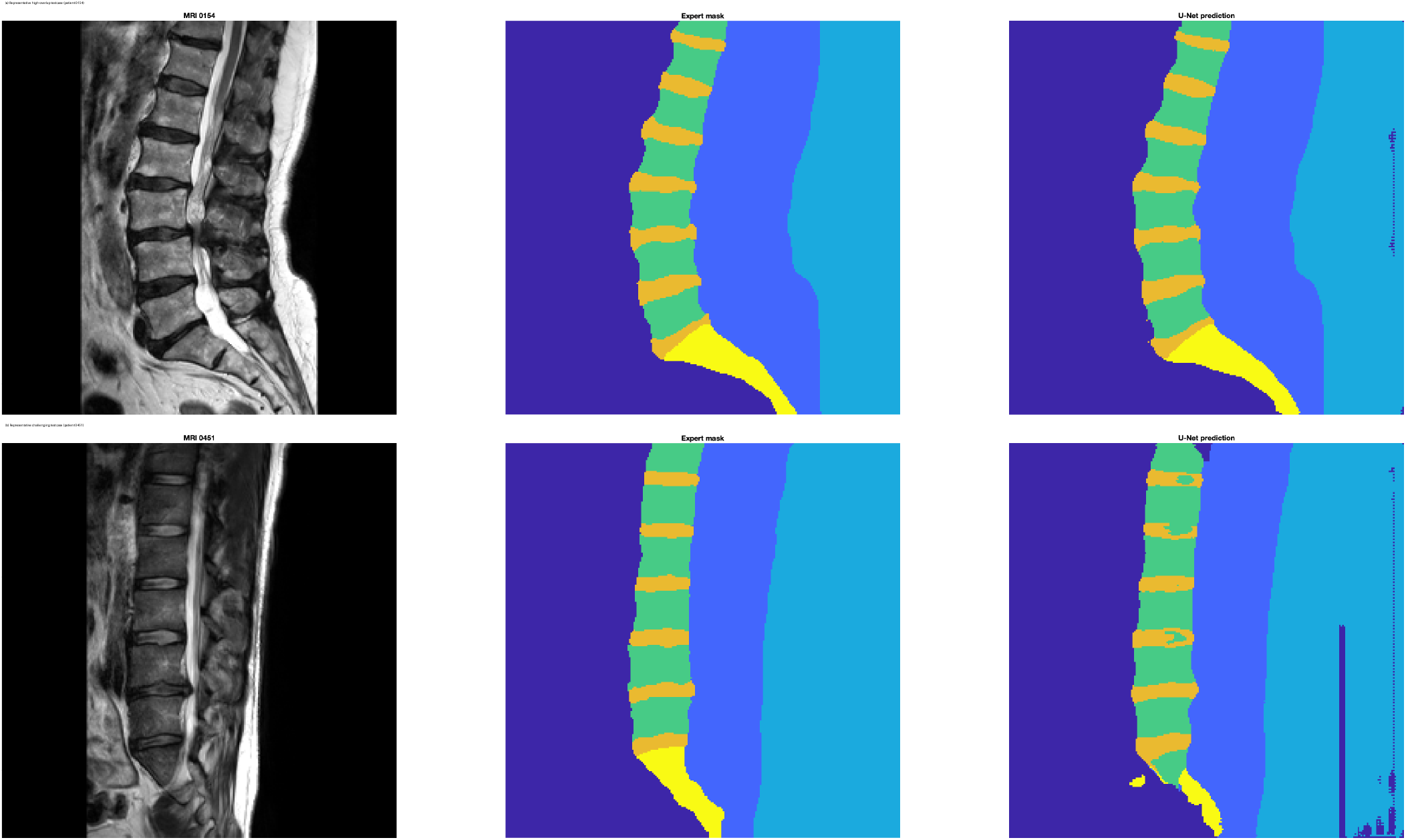
Representative medical MRI segmentation results derived from the public LSS MRI AISSLab dataset. Each row shows sagittal T2-weighted MRI, expert-refined reference mask, and frozen U-Net prediction. (a) A high-overlap test case. (b) The lowest patient-mean-Dice test case, included deliberately to show a failure/challenging example rather than only favorable images. Dataset images are used with attribution to [9], [10]; predictions are from the present study.

**Fig. 5.**
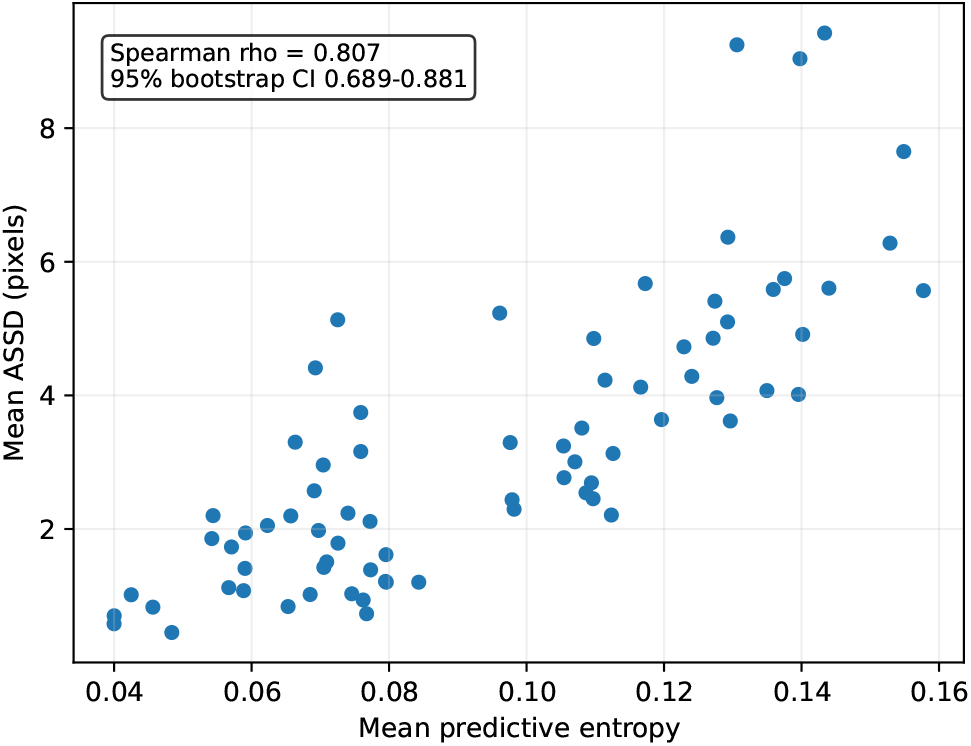
Patient-level association between mean segmentation predictive entropy and mean ASSD on the internal test set. The strong monotonic association supports entropy as a useful quality-control indicator for this model.

**Fig. 6.**
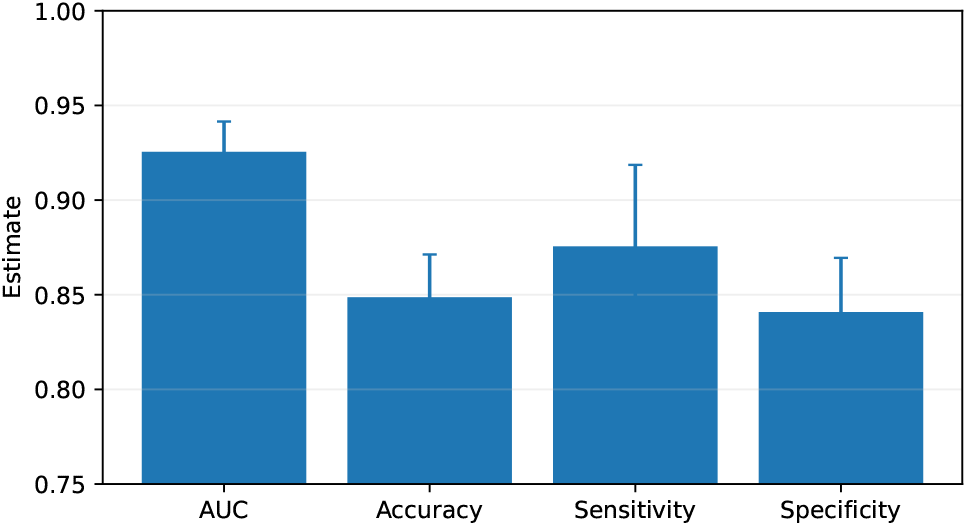
Held-out 2.5-D performance at the tuning-locked threshold. Error bars denote patient-clustered bootstrap 95% CIs.

**Fig. 7.**
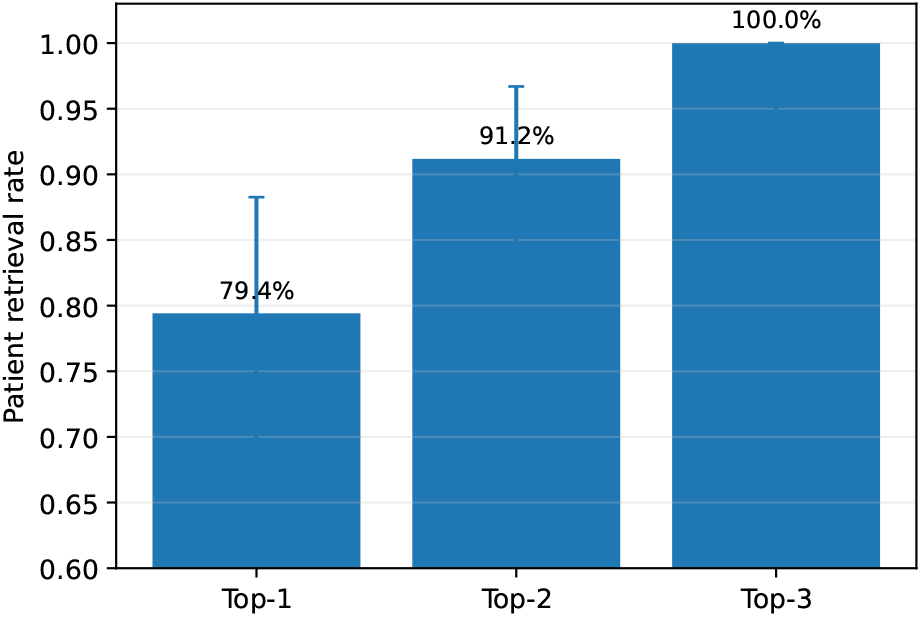
Patient-level top-*k* slice retrieval. Exact binomial intervals are used because top-3 achieved 68/68 hits.

### C. Segmentation Entropy Tracked Surface Error

Mean predictive entropy was weakly and inversely associated with patient-mean Dice (*ρ* = −0.236, bootstrap CI −0.484 to 0.037) but more strongly associated with surface errors. Mean entropy correlated with mean HD95 at *ρ* = 0.547 (0.349–0.713) and with mean ASSD at *ρ* = 0.807 (0.689– 0.881). The strong entropy–ASSD association suggests that the probability maps contain useful information about where segmentation boundaries are less reliable, although this does not constitute prospective calibration.

### D. 2.5-D Slice Ranking Generalized at a Locked Operating Point

The 2-D and 2.5-D selectors achieved tuning AUCs of 0.931 and 0.933, respectively; the 2.5-D model was selected. On 998 internal-test slices (225 annotation-bearing, 773 annotation-negative), AUC was 0.926 (95% patient-clustered CI, 0.909– 0.941). The tuning-only threshold satisfying sensitivity ≥ 0.90 was 0.114394. Applied unchanged to the test set, it achieved accuracy 0.849 (0.826–0.871), sensitivity 0.876 (0.832–0.919), and specificity 0.841 (0.813–0.869). This is materially different from the arbitrary 0.5 threshold, which gave high specificity but low sensitivity; the locked analysis demonstrates a clinically more relevant operating point without test-set tuning.

Patient ranking was even more operationally interpretable. Among 68 test patients with at least one annotated slice, top-1 recovery was 54/68 (79.4%; exact 95% CI 67.9–88.3%), top-2 was 62/68 (91.2%; 81.8–96.7%), and top-3 was 68/68 (100%; 94.7–100%). The median rank of the first annotation-bearing slice was 1, the 90th percentile was 2, and the maximum was 3.

**TABLE IV.** Frozen 2.5-D Slice-Selection Test Performance.

| Metric | Estimate | 95% CI |
| --- | --- | --- |
| ROC AUC | 0.926 | 0.909–0.941 |
| Accuracy | 0.849 | 0.826–0.871 |
| Sensitivity | 0.876 | 0.832–0.919 |
| Specificity | 0.841 | 0.813–0.869 |

**Table V.** Detector Performance on the Internal Test Set.

| Evaluation | IoU | AP | Recall / Precision |
| --- | --- | --- | --- |
| Localization | 0.50 | 0.530 | 0.754 / 0.518 |
| Localization | 0.75 | 0.046 | 0.188 / 0.129 |
| Class-aware | 0.50 | 0.345 | 0.567 / 0.389 |
| Class-aware | 0.75 | 0.030 | 0.144 / 0.099 |

### E. Precise Foraminal Localization Was the Main Bottleneck

On 225 annotation-bearing test slices, the detector produced 676 predictions for 464 reference boxes, with 350 one-to-one matches at IoU ≥ 0.50. Localization AP50 was 0.530, recall 0.754, and terminal precision 0.518. At the stricter IoU 0.75, AP fell to 0.046 and recall to 0.188. Requiring the severity class to match reduced AP50 to 0.345 and AP75 to 0.030.

The annotation-negative audit provided a second view of detector behavior. Across 773 XML-negative test slices, mean predictions per slice were 0.529 (95% patient-clustered CI 0.464–0.590); 29.1% (26.0–32.0%) had at least one prediction and 14.5% (12.4–16.6%) had at least two. This does not equal clinical false-positive rate because unannotated slices can contain anatomy or subtle pathology not represented by XML. It does, however, show that the current detector is better viewed as a candidate generator than a precise stand-alone locator.

### F. Model Selection Avoided Majority-Class Collapse

The severity tuning set was strongly imbalanced, so model selection based only on raw accuracy would have been misleading. SqueezeNet transfer learning achieved tuning accuracy 0.642 versus 0.617 for the scratch CNN, but its balanced accuracy was 0.250, macro-F1 0.195, and QWK 0.000, indicating near-complete majority-class collapse. The scratch CNN instead achieved tuning balanced accuracy 0.489, macro-F1 0.466, and QWK 0.572 and was frozen for testing.

### G. Localized Severity Grading Preserved Ordinal Information

On 464 expert-defined test ROIs from 68 patients, the scratch CNN achieved accuracy 0.649 (95% CI 0.597– 0.699), balanced accuracy 0.519 (0.457–0.578), macro-F1 0.493 (0.436–0.545), MAE 0.440 grades (0.377–0.505), QWK 0.638 (0.550–0.706), and within-one-grade agreement 0.927 (0.904–0.948). The majority-class baseline is an important comparator: because 326/464 test ROIs were normal, an all-normal classifier would achieve 70.3% raw accuracy while having no useful discrimination among abnormal grades. Accordingly, QWK, balanced accuracy, macro-F1, and ordinal AUC are more informative than raw accuracy.

Class-specific recall was 0.752 for normal, 0.300 for mild, 0.469 for moderate, and 0.556 for severe. Severe classification therefore remained imperfect but was not collapsed into the majority class. The confusion matrix (Fig. 9) shows that most errors occurred between neighboring grades; only 7.3% of test predictions differed by two or more grades.

Continuous probabilities gave stronger clinically interpretable discrimination than nominal accuracy. AUC was 0.856 (0.815–0.894) for any stenosis, 0.893 (0.850–0.932) for moderate-or-worse disease, and 0.912 (0.868–0.951) for severe disease. One-vs-rest grade-1 AUC was the weakest at 0.686, consistent with ambiguity between mild and neighboring grades.

**TABLE VI.** Four-Grade CNN Test Performance.

| Metric | Estimate | 95% CI |
| --- | --- | --- |
| Accuracy | 0.649 | 0.597–0.699 |
| Balanced accuracy | 0.519 | 0.457–0.578 |
| Macro-F1 | 0.493 | 0.436–0.545 |
| Ordinal MAE | 0.440 | 0.377–0.505 |
| QWK | 0.638 | 0.550–0.706 |
| Within one grade | 0.927 | 0.904–0.948 |

**TABLE VII.** Ordinal-Boundary Severity Discrimination.

| Endpoint | AUC | 95% CI | AP |
| --- | --- | --- | --- |
| Any stenosis ($\geq 1$) | 0.856 | 0.815–0.894 | 0.754 |
| Moderate-or-worse ($\geq 2$) | 0.893 | 0.850–0.932 | 0.685 |
| Severe ($\geq 3$) | 0.912 | 0.868–0.951 | 0.608 |

### H. Medical ROI Examples and Qualitative Occlusion Sensitivity

Figure 11 provides direct MRI examples rather than only summary statistics. These examples are correctly classified normal and mild ROIs generated during the frozen explainability stage. The strongest occlusion-response regions overlap the central foraminal morphology in both examples. This observation is qualitative; the heat maps do not establish causal reasoning, do not validate clinical saliency, and should not be used as a stand-alone explanation of model behavior.

### I. CNN and Radiomics Had Similar Accuracy but Very Different Ordinal Quality

The radiomics-SVM baseline achieved accuracy 0.647, nearly identical to the CNN’s 0.649. Accuracy alone would therefore imply equivalence. Patient-clustered paired analysis contradicted that conclusion: the CNN improved balanced accuracy by 0.205 (95% CI 0.132–0.276), macro-F1 by 0.173 (0.100–0.240), QWK by 0.360 (0.226–0.487), and within-one-grade agreement by 0.058 (0.020–0.097). Ordinal MAE decreased by 0.103 grades (paired CNN-minus-radiomics difference −0.103, CI −0.204 to − 0.004). The raw accuracy difference was only 0.002 (CI −0.056 to 0.058).

These paired intervals show that learned local image features preserved substantially more minority-class and ordinal information than the 29-feature handcrafted baseline even though conventional accuracy was almost the same.

### J. Whole-Volume 3-D Classification Was Weak and Poorly Calibrated

The whole-volume 3-D CNN was evaluated on 68 patients (35 positive, 33 negative). Test AUC was 0.639 (95% CI 0.496–0.765), with the interval crossing 0.5. At threshold 0.5, sensitivity was 0.971 but specificity only 0.061. Mean model score was 0.864 in positive patients and 0.800 in negative patients. Brier score was 0.340 and 10-bin ECE was 0.331, demonstrating substantial probability miscalibration.

Scanner-subgroup AUCs were 0.654 for Philips Ingenia 1.5 T (*n* = 33), 0.864 for GE Signa HDxt 1.5 T (*n* = 18), and 0.609 for Philips Ingenia 3 T (*n* = 16). These subgroups are too small for scanner-specific claims, especially because specificity was extremely poor. The apparently high GE subgroup AUC should therefore be interpreted as descriptive instability rather than evidence of scanner superiority.

### K. Exploratory Level and Laterality Analysis

Severity performance varied across lumbar levels, but the analysis is descriptive because class prevalence changes strongly by level. Moderate-or-worse AUCs were 0.841 at L1– L2, 0.784 at L2–L3, 0.921 at L3–L4, 0.870 at L4–L5, and 0.691 at L5–S1. QWK was highest at L3–L4 (0.705) and L4– L5 (0.647) and lowest at L5–S1 (0.172). Left-sided ROIs had QWK 0.704 versus 0.537 on the right, with moderate-or-worse AUC 0.912 versus 0.862. These patterns may reflect differing case mix, visibility, or annotation frequency rather than true biological laterality and require independent replication.

## V. Discussion

### A. Principal Finding: Localization and Spatial Constraint Matter

The central result is not that every stage performed well. The central result is that performance changed systematically with spatial constraint. Mid-sagittal segmentation was highly accurate, 2.5-D slice ranking reliably reduced the search space, and expert-localized ROI grading preserved useful ordinal information. In contrast, precise object localization remained difficult and an unconstrained whole-volume 3-D classifier was weak and poorly calibrated. This pattern supports a modular strategy in which anatomical and slice-level localization precede severity inference rather than expecting one compact classifier to discover all relevant structure automatically.

This conclusion is stronger than reporting only the best model because the weaker components were intentionally retained. The 3-D AUC confidence interval crossing 0.5 and the detector’s AP75 of 0.046 are negative results in the conventional sense, but they identify where information is lost. In a clinically oriented engineering study, identifying a failure boundary is valuable: it discourages premature claims of end-to-end automation and directs future work toward precise localization and level-aware modeling.

### B. Clinical Meaning of the Slice-Selection Result

The slice selector is one of the clearest successes. An AUC of 0.926 demonstrates strong ranking, but AUC alone does not specify a clinically usable operating point. The initial 0.5 threshold had high specificity and low sensitivity; after locking a threshold on tuning data, the internal test sensitivity/specificity became 0.876/0.841 without any test-set optimization. This distinction is important because many AI studies report threshold-dependent measures without explaining how the threshold was chosen.

**Fig. 8.**
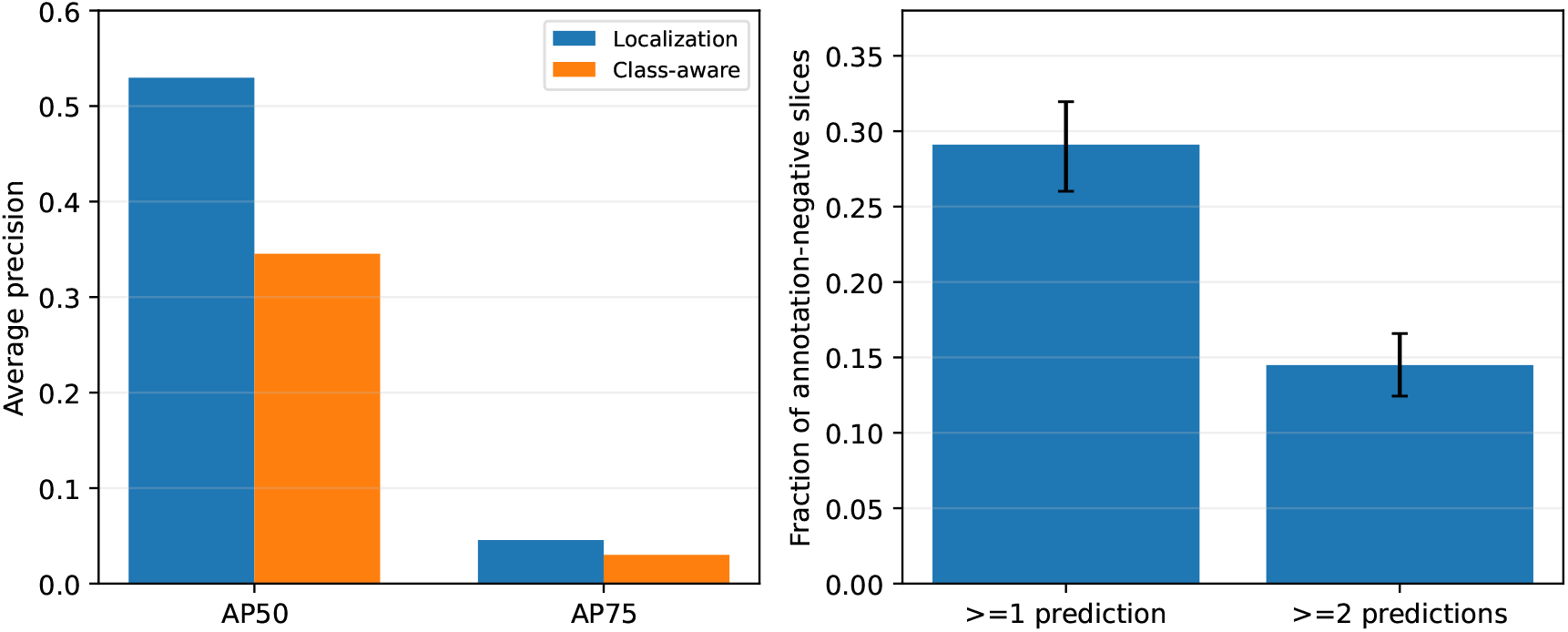
Detector failure analysis. Left: localization and class-aware AP deteriorate sharply from IoU 0.50 to 0.75, indicating imprecise boxes despite moderate coarse localization. Right: prediction burden on 773 annotation-negative test slices using the same frozen inference thresholds.

**Fig. 9.**
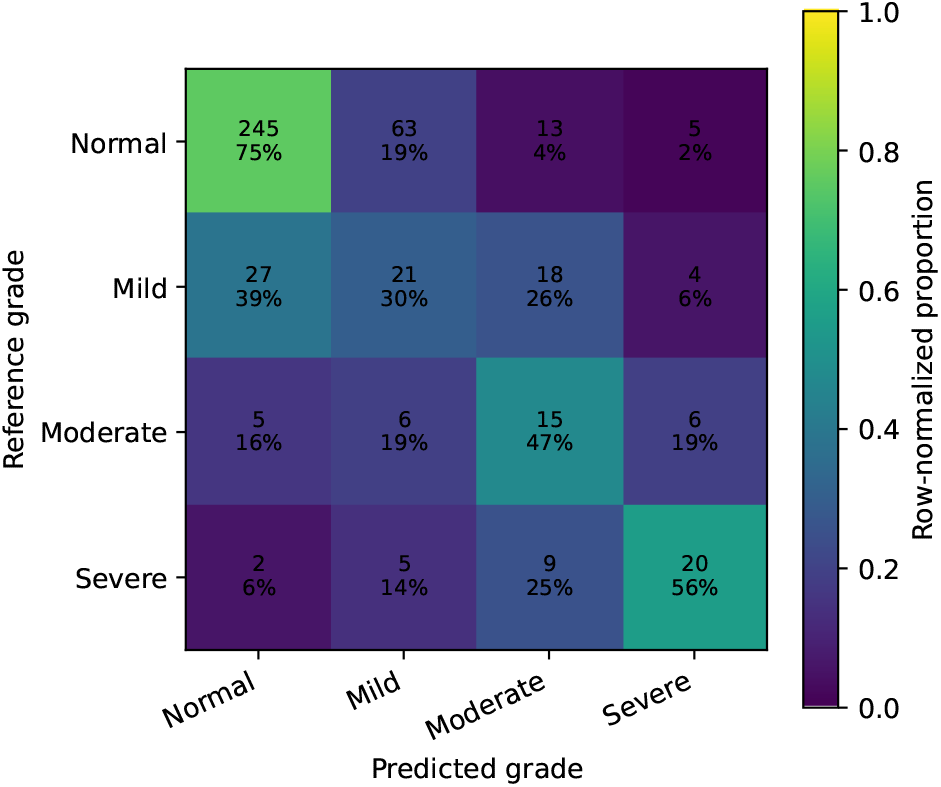
Held-out severity confusion matrix. Percentages are normalized within each reference grade. The error structure is predominantly ordinal/adjacent rather than arbitrary.

**Fig. 10.**
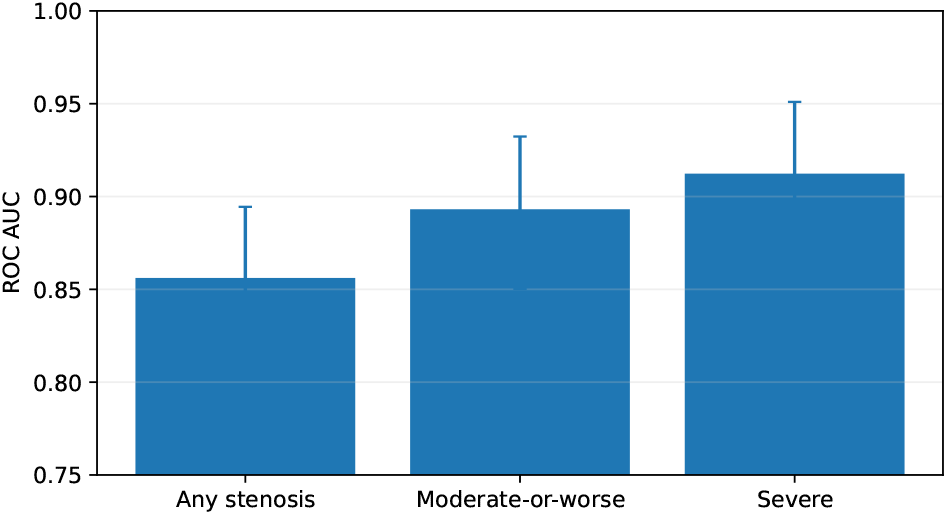
ROC AUC for clinically meaningful ordinal severity boundaries derived from frozen four-class probabilities.

Top-*k* retrieval offers a different workflow interpretation. The top three slices recovered at least one annotation-bearing slice in all 68 positive test patients, with a lower exact confidence bound of 94.7%. This does not mean all stenotic levels were recovered, nor that a radiologist could review only three slices in clinical practice. It does suggest that a ranking model can substantially narrow the search space for downstream candidate generation while preserving at least one annotated slice per patient in this internal test set.

### C. Why Detection Was Harder Than Ranking

Slice ranking asks whether a slice contains the broad visual context associated with foraminal annotations. Detection asks for precise spatial coordinates of small lateral structures. The severe AP drop from IoU 0.50 to 0.75 therefore has an anatomically plausible interpretation: the detector often found approximately relevant regions without matching expert boxes tightly. The negative-slice audit further shows that the network generates candidates on a nontrivial fraction of unannotated slices.

These results argue for improving the localization stage before claiming end-to-end grading. Promising directions include explicit hard-negative sampling, feature pyramids, level-aware anatomical priors, bilateral symmetry constraints, and a detector objective designed for small elongated anatomical regions. The segmentation stage may provide a useful coordinate system for such priors even though the current mid-sagittal masks and lateral foramina are not spatially identical targets.

### D. Ordinal Grading Is Better Evaluated by Agreement Than Accuracy

The severity experiment demonstrates a broader methodological point. A classifier predicting every ROI as normal would achieve 70.3% test accuracy, greater than the selected CNN’s 64.9%. If accuracy were the sole endpoint, the learned model would appear inferior to a trivial rule. That conclusion would be clinically and statistically incomplete because the trivial model cannot grade disease.

**Fig. 11.**
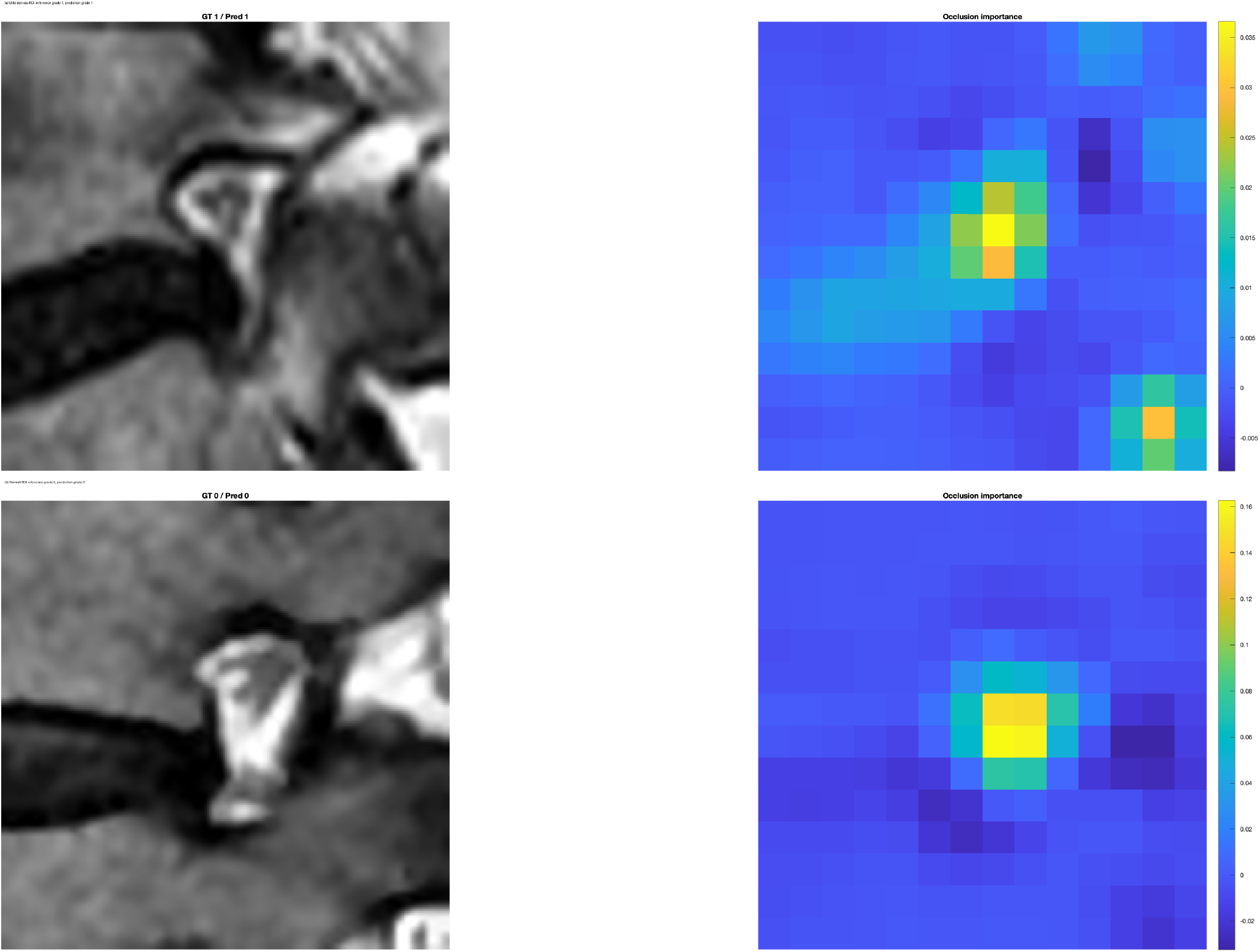
Representative expert-defined foraminal MRI ROIs and occlusion-sensitivity maps from the frozen severity CNN. (a) Correctly classified mild ROI (reference 1, predicted 1). (b) Correctly classified normal ROI (reference 0, predicted 0). These examples demonstrate the actual medical-image inputs and model sensitivity pattern; they are illustrative rather than quantitative evidence. Dataset-derived images are attributed to [9], [10].

**Fig. 12.**
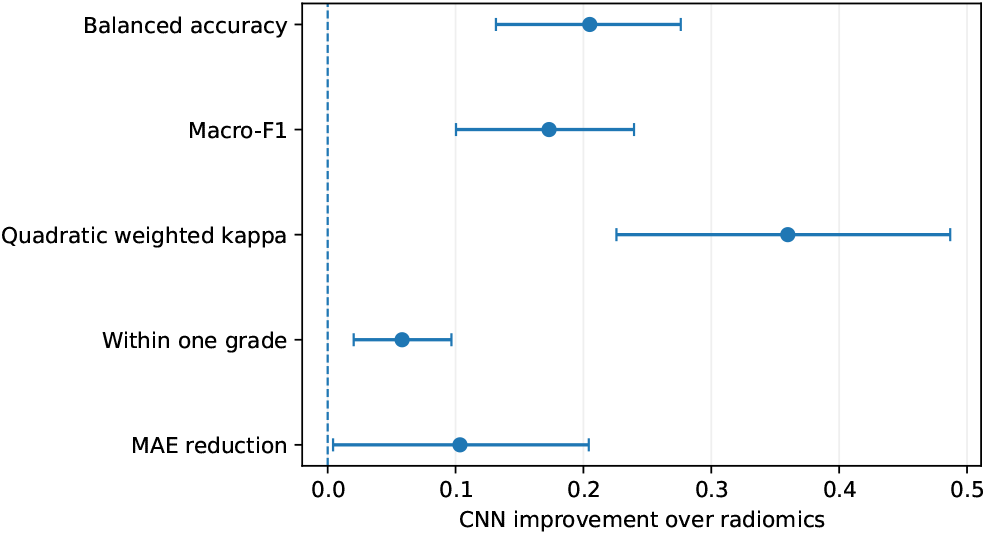
Paired patient-bootstrap improvements of the scratch CNN over the radiomics-SVM baseline. Positive values favor the CNN; for MAE the plotted quantity is MAE reduction.

**Fig. 13.**
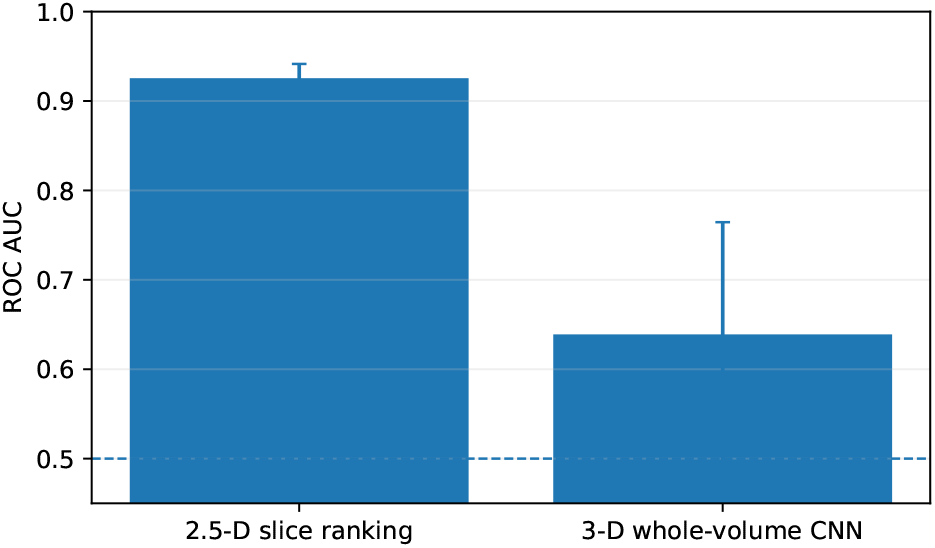
Performance at two spatial scales. The endpoints differ, so the bars are not a formal paired comparison. The figure illustrates the strong informative-slice signal versus the weak exploratory whole-volume classifier.

The CNN instead achieved QWK 0.638, 92.7% within-one-grade agreement, and moderate-or-worse AUC 0.893. It also materially outperformed radiomics on balanced accuracy, macro-F1, QWK, and ordinal MAE despite essentially identical raw accuracy. The comparison demonstrates why class-balanced and ordinal endpoints are essential for imbalanced severity tasks. Grade 1 remained the most difficult class, which is consistent with the morphological continuum between normal/mild and mild/moderate disease.

**Fig. 14.**
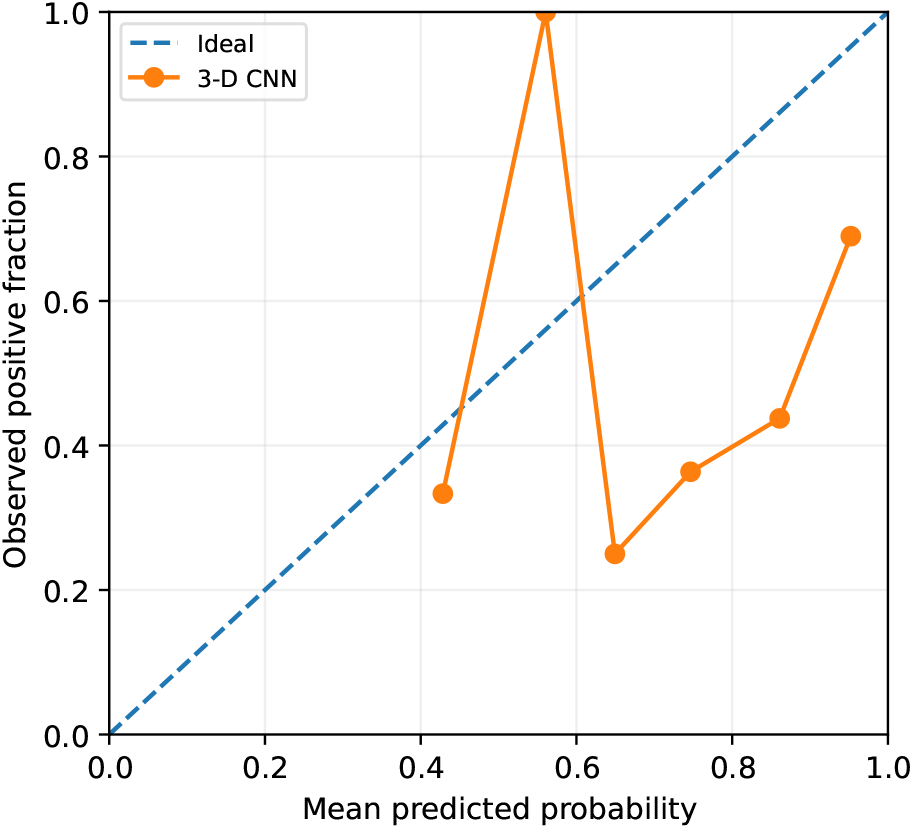
Exploratory 3-D CNN calibration. Predicted probabilities were concentrated at high values for both classes, consistent with the low specificity, Brier score 0.340, and ECE 0.331.

### E. Relationship to Prior Work

The current results should not be read as a direct numerical competition with studies using different series, labels, reader standards, or test definitions. Hallinan *et al*. achieved high agreement for dichotomized neural foraminal stenosis with a two-stage system and external testing, but used sagittal T1 and axial T2 MRI from an institutional cohort with expert radiologist labels [7]. That work represents a higher level of clinical validation than the present public-dataset internal test and is therefore an important benchmark rather than a target to be “beaten.”

On the same AISSLab resource, the data descriptor reported successful component-level CAD development and strong anatomical segmentation [9]. Phadke *et al*. focused on ordinal grading of expert-defined ROIs and fused deep features with interpretable morphometry; automated full-exam localization was outside their scope [11]. The present study is complementary: it asks how information changes across spatial scales under one frozen split, performs a validation-locked slice-selection analysis, quantifies annotation-negative detector behavior, compares deep and radiomic ROI representations on identical cases, and retains a negative whole-volume result.

### F. Uncertainty as a Quality-Control Signal

A useful quality-control measure should correlate with error. The strong entropy–ASSD correlation (*ρ* = 0.807) is therefore more informative than displaying entropy maps alone. Mean entropy was less strongly related to Dice, which is plausible because Dice measures bulk overlap whereas ASSD is sensitive to boundary displacement. This suggests that probability entropy may be particularly useful for identifying boundary uncertainty. However, the analysis is post hoc and internal; it should not be interpreted as a calibrated guarantee of segmentation correctness.

### G. Reproducibility and Computational Provenance

Complex computational imaging studies require a transparent provenance chain so that model development, frozen evaluation, and publication postprocessing cannot be inadvertently conflated. The present work addresses this requirement through deterministic patient splitting, stage-level completion locks, frozen test predictions, validation-only threshold selection, patient-clustered resampling, paired comparisons on identical test ROIs, retention of unfavorable results, and explicit publication postprocessing scripts. Qualitative figures include a deliberately challenging segmentation case rather than only visually successful examples. These measures do not replace independent external replication, but they make the computational chain auditable and reduce the opportunity for accidental leakage or selective reporting.

## VI. Limitations

This study has important limitations that constrain the conclusions.

First, the internal test set comes from the same public dataset as training/tuning. Although scanners and field strengths are heterogeneous, this is not external testing. Generalization to other institutions, vendors, MRI protocols, populations, and annotation practices remains unknown.

Second, the ROI severity CNN was evaluated on expert-defined boxes. Its QWK and ordinal AUC therefore characterize *grading conditional on correct localization*, not end-to-end detector-to-grader performance. The detector is currently the major bottleneck, as shown by low AP75 and nontrivial prediction burden on annotation-negative slices.

Third, XML-negative slices are not clinically normal controls. Some may contain unannotated normal foramina or abnormalities not selected by the source annotation process. The 29.1% prediction rate is therefore a prediction-burden measure, not patient-level clinical specificity.

Fourth, the segmentation target is the mid-sagittal slice, whereas foraminal disease is lateral. The excellent segmentation performance should not be interpreted as direct segmentation of the neural foramen. Its main relevance is anatomical normalization, quality control, and potential future spatial priors.

Fifth, the whole-volume comparator was deliberately compact. Its poor result demonstrates that this implementation did not solve patient-level disease discovery; it does not establish that all modern 3-D architectures or vision transformers would fail.

Sixth, no external clinical outcomes, symptoms, surgical decisions, or reader-study comparisons are available in the current experiment. Imaging grade should not be equated with pain or treatment need [2], [3].

Seventh, class prevalence differs markedly by lumbar level, and subgroup sample sizes are small. The level/laterality analyses are hypothesis-generating only.

Finally, repeated methodological development on a single public resource can create researcher degrees of freedom even when the test set is protected. The strongest next step is independent external testing using a separately curated lumbar MRI cohort and a locked end-to-end pipeline.

## VII. Future Work

The next technical priorities follow directly from the failure analysis. First, the detector should be redesigned around hard-negative training, multi-scale features, and anatomy-conditioned priors. Second, end-to-end evaluation should propagate detector uncertainty into ROI grading rather than substituting expert boxes. Third, severity modeling should use an explicitly ordinal loss or cumulative-link formulation and compare against the present class-weighted nominal CNN. Fourth, scanner and level robustness should be evaluated on larger external cohorts. Fifth, a prospective reader-assistance study should measure whether slice ranking and candidate localization reduce interpretation time without increasing clinically important misses. Finally, segmentation entropy could be used to trigger automatic quality-control review if its error association replicates externally.

## VII. Conclusion

A comprehensive leakage-controlled analysis of 500 multi-scanner sagittal lumbar MRI examinations showed that *where the model is asked to look* is as important as the classifier itself. Anatomical segmentation and 2.5-D slice ranking were strong; a validation-locked slice threshold retained high sensitivity and specificity, and the top three ranked slices recovered an annotation-bearing slice in all 68 positive test patients. Conditional on expert localization, the severity CNN preserved meaningful ordinal information and materially outperformed handcrafted radiomics on class-balanced and weighted-agreement measures. In contrast, precise bounding-box localization remained difficult and whole-volume 3-D classification was weak and poorly calibrated. These findings support a modular localization-aware architecture for lumbar foraminal stenosis AI and provide a transparent map of both capability and failure modes. External testing and end-to-end detector-to-grader evaluation are required before any clinical deployment claim is justified.

## Data Availability

The LSS MRI AISSLab dataset is publicly available through Mendeley Data, Version 4, DOI 10.17632/rgb77xm3jf.4. The dataset page provides the current license terms. No raw patient images are redistributed with this manuscript beyond derived illustrative figures used to communicate the reported results.

https://data.mendeley.com/datasets/rgb77xm3jf/4

## Data Availability

The LSS MRI AISSLab dataset is publicly available through Mendeley Data, Version 4, DOI 10.17632/rgb77xm3jf.4 [10]. The dataset page identifies the resource for scientific research and provides the current license terms. No raw patient images are redistributed in the source package of this manuscript beyond derived illustrative figures required to communicate the reported results.

## Code and Reproducibility Availability

The complete experiment was implemented in MATLAB R2024b. The submission package contains the LaTeX source, frozen-result tables, publication figures, and CSV-only post-processing code used to derive confidence intervals, threshold-locked performance, top-*k* retrieval, uncertainty/error association, severity subgroup summaries, and 3-D calibration. These scripts do not retrain models or tune parameters on the test set. The complete training pipeline is archived by the author and can be supplied to reviewers or released in a public repository for final publication.

## Author Contributions

Y. Riyazifar performed the study design, software implementation, computational experiments, statistical analysis, visualization, interpretation, quality-control audit, and manuscript preparation.

## Appendix A Extended Quantitative Results

### A. Severity Class-Specific Performance

Table VIII shows the class-specific behavior of the selected CNN. Mild stenosis was the most difficult class, with recall 0.300 and F1 0.255, whereas normal and severe classes were better separated. This pattern explains why macro-F1 is materially lower than raw accuracy.

**TABLE V111.**
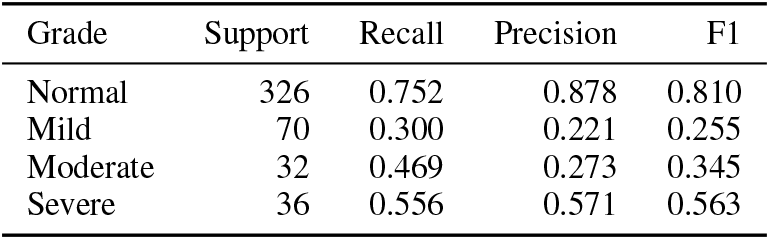
Class-Specific Test Performance of the Severity CNN.

### B. One-vs-Rest Severity Probability Performance

**TABLE IX.**
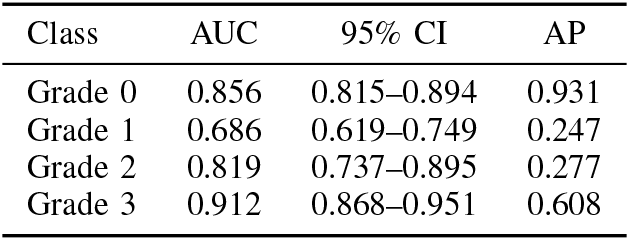
One-vs-Rest Severity AUC and Average Precision.

### C. Per-Grade Detector Performance

Class-aware AP50 was strongly dominated by grade 0. Per-grade AP50 values were 0.490 (grade 0), 0.033 (grade 1), 0.014 (grade 2), and 0.130 (grade 3). These values reinforce that the detector’s classification head is not sufficiently reliable for direct clinical grading and justify evaluating severity separately on expert-defined ROIs.

## Appendix B Exploratory Severity Subgroups

The subgroup analyses below were prespecified as descriptive publication postprocessing of frozen test predictions and were not used for model selection or threshold tuning. Because grade prevalence and sample size vary by level and side, these results are hypothesis-generating rather than evidence of biological or scanner-independent subgroup effects. The complete subgroup metrics are retained here to make the observed heterogeneity auditable rather than selectively reporting only favorable strata.

**TABLE X.** Descriptive Severity CNN Agreement and Discrimination by Lumbar Level.

| Level | ROIs | Patients | QWK | AUC $\geq 2$ |
| --- | --- | --- | --- | --- |
| L1–L2 | 83 | 50 | 0.275 | 0.841 |
| L2–L3 | 105 | 59 | 0.233 | 0.784 |
| L3–L4 | 126 | 63 | 0.705 | 0.921 |
| L4–L5 | 106 | 56 | 0.647 | 0.870 |
| L5–S1 | 44 | 30 | 0.172 | 0.691 |

**TABLE XI.** Descriptive Class-Balanced and Ordinal Error Metrics by Lumbar Level.

| Level | Macro-F1 | Bal. Acc. | MAE | Within-1 |
| --- | --- | --- | --- | --- |
| L1–L2 | 0.268 | 0.382 | 0.145 | 1.000 |
| L2–L3 | 0.303 | 0.432 | 0.295 | 0.943 |
| L3–L4 | 0.461 | 0.470 | 0.405 | 0.952 |
| L4–L5 | 0.487 | 0.502 | 0.632 | 0.896 |
| L5–S1 | 0.337 | 0.420 | 0.977 | 0.750 |

**TABLE XII.** Descriptive Severity CNN Performance by Laterality.

| Side | ROIs | Patients | QWK | Macro-F1 | Bal. Acc. |
| --- | --- | --- | --- | --- | --- |
| Left | 246 | 64 | 0.704 | 0.573 | 0.609 |
| Right | 218 | 67 | 0.537 | 0.364 | 0.384 |

**TABLE XIII.** Descriptive Laterality Error and Discrimination Metrics.

| Side | MAE | Within-1 | AUC $\geq 2$ |
| --- | --- | --- | --- |
| Left | 0.402 | 0.939 | 0.912 |
| Right | 0.482 | 0.913 | 0.862 |

## Appendix C Reproducibility and Audit Notes

The final V8.1.11 run completed all analysis stages before publication postprocessing. The postprocessing sequence intentionally separated model training from publication statistics. Version 1 calculated patient-bootstrap severity/radiomics comparisons, detector AP, radiomics PCA dimensionality, validation-locked slice thresholds, and detector prediction burden on annotation-negative test slices. Version 2 added patient-clustered CIs for the locked slice endpoint and exact top-*k* patient retrieval intervals. Version 3 added frozen CSV-only uncertainty/error association, descriptive severity subgroups, and 3-D calibration. None of these scripts changed learned weights, data partitions, or test thresholds.

The following safeguards were used:

- patient identity, not image identity, defined data splitting;
- the same patient partition was inherited by slices and ROIs;
- architecture/model selection used the tuning set;
- the slice decision threshold was selected on tuning predictions only;
- paired CNN/radiomics intervals resampled identical patients;
- unfavorable detector and 3-D results were retained;
- qualitative segmentation includes a challenging low-Dice case;
- “annotation-negative” is distinguished from “clinically normal”;
- no external-testing claim is made.

## Appendix D Reporting Checklist Alignment

The manuscript was structured around CLAIM 2024 principles [12]: the title and abstract identify the AI methodology and public data source; study design and patient-level splitting are explicit; the reference standard and source ethics are described; training/tuning/test roles are separated; preprocessing and task definitions are reported; uncertainty and failure analysis are provided; negative results are retained; and intended use is framed as research/decision support rather than autonomous diagnosis. Items requiring future external data, prospective evaluation, clinical impact analysis, or a reader study are explicitly identified as limitations rather than implied to have been completed.

## References

[1] N. Mamisch, M. Brumann, J. Hodler, U. Held, F. Brunner, and J. Steurer, “Radiologic criteria for the diagnosis of spinal stenosis: Results of a Delphi survey,” Radiology, vol. 264, no. 1, pp. 174–179, 2012, doi: 10.1148/radiol.12111930.

[2] H. J. Park, S. S. Kim, S.-Y. Lee, N.-H. Park, M.-H. Rho, H.-P. Hong, H.-J. Kwag, S.-H. Kook, and S.-H. Choi, “Clinical correlation of a new MR imaging method for assessing lumbar foraminal stenosis,” AJNR Am. J. Neuroradiol., vol. 33, no. 5, pp. 818–822, 2012, doi: 10.3174/ajnr.A2870.

[3] E. Sartoretti, M. Wyss, A. Alfieri, C. A. Binkert, C. Erne, S. Sartoretti-Schefer, and T. Sartoretti, “Introduction and reproducibility of an updated practical grading system for lumbar foraminal stenosis based on high-resolution MR imaging,” Sci. Rep., vol. 11, Art. no. 12000, 2021, doi: 10.1038/s41598-021-91462-2.

[4] S. Lee, J. W. Lee, J. S. Yeom, K.-J. Kim, H.-J. Kim, S. K. Chung, and H. S. Kang, “A practical MRI grading system for lumbar foraminal stenosis,” AJR Am. J. Roentgenol., vol. 194, no. 4, pp. 1095–1098, 2010, doi: 10.2214/AJR.09.2772.

[5] A. Jamaludin, T. Kadir, and A. Zisserman, “SpineNet: Automated classification and evidence visualization in spinal MRIs,” Med. Image Anal., vol. 41, pp. 63–73, 2017, doi: 10.1016/j.media.2017.07.002.

[6] J.-T. Lu, S. Pedemonte, B. Bizzo, S. Doyle, K. P. Andriole, M. H. Michalski, R. G. Gonzalez, and S. R. Pomerantz, “Deep Spine: Automated lumbar vertebral segmentation, disc-level designation, and spinal stenosis grading using deep learning,” in Proc. 3rd Mach. Learn. Healthcare Conf., PMLR, vol. 85, 2018, pp. 403–419.

[7] J. T. P. D. Hallinan et al., “Deep learning model for automated detection and classification of central canal, lateral recess, and neural foraminal stenosis at lumbar spine MRI,” Radiology, vol. 300, no. 1, pp. 130–138, 2021, doi: 10.1148/radiol.2021204289.

[8] M. A. Al-Antari et al., “Evaluating AI-powered predictive solutions for MRI in lumbar spinal stenosis: A systematic review,” Artif. Intell. Rev., vol. 58, Art. no. 221, 2025, doi: 10.1007/s10462-025-11185-y.

[9] O. F. Abdulmahmod et al., “Medical spine sagittal MRI dataset for segmentation and foraminal stenosis detection,” Sci. Data, vol. 13, Art. no. 809, 2026, doi: 10.1038/s41597-026-07138-x.

[10] O. F. Abdulmahmod et al., “LSS MRI AISSLab Dataset: Medical Spine Sagittal MRI Dataset for Segmentation and Foraminal Stenosis Detection,” Mendeley Data, Version 4, 2026, doi: 10.17632/rgb77xm3jf.4.

[11] R. A. Phadke et al., “Ordinal deep learning for lumbar foraminal stenosis grading on sagittal MRI,” J. Imaging, vol. 12, no. 8, Art. no. 388, 2026, doi: 10.3390/jimaging12080388.

[12] A. S. Tejani et al., “Checklist for Artificial Intelligence in Medical Imaging (CLAIM): 2024 update,” Radiol. Artif. Intell., vol. 6, no. 4, Art. no. e240300, 2024, doi: 10.1148/ryai.240300.

[13] O. Ronneberger, P. Fischer, and T. Brox, “U-Net: Convolutional networks for biomedical image segmentation,” in Medical Image Computing and Computer-Assisted Intervention–MICCAI 2015, LNCS 9351, 2015, pp. 234–241, doi: 10.1007/978-3-319-24574-428.

[14] K. He, X. Zhang, S. Ren, and J. Sun, “Deep residual learning for image recognition,” in Proc. IEEE Conf. Comput. Vis. Pattern Recognit. (CVPR), 2016, pp. 770–778, doi: 10.1109/CVPR.2016.90.

[15] O. Oktay et al., “Attention U-Net: Learning where to look for the pancreas,” arXiv:1804.03999, 2018, doi: 10.48550/arXiv.1804.03999.

[16] F. N. Iandola, S. Han, M. W. Moskewicz, K. Ashraf, W. J. Dally, and K. Keutzer, “SqueezeNet: AlexNet-level accuracy with 50x fewer parameters and <0.5 MB model size,” arXiv:1602.07360, 2016.

